# The ratio between SARS-CoV-2 RNA viral load and culturable viral titer differs depending on stage of infection

**DOI:** 10.1101/2023.07.06.23292300

**Authors:** Michael K. Porter, Alexander Viloria Winnett, Linhui Hao, Natasha Shelby, Jessica A. Reyes, Noah W. Schlenker, Anna E. Romano, Colton Tognazzini, Matthew Feaster, Ying-Ying Goh, Michael Gale, Rustem F. Ismagilov

**Affiliations:** Division of Chemistry and Chemical Engineering, California Institute of Technology, Pasadena CA 91125; Department of Immunology, University of Washington, Seattle WA 98109; Pasadena Public Health Department, Pasadena CA 91125

**Keywords:** SARS-CoV-2, incident, culture, infectivity, longitudinal

## Abstract

Analysis of incident, longitudinal RNA viral loads in saliva and nasal swabs and culturable viral titers in nasal swabs collected twice-daily by a tricenarian male infected with SARS-CoV-2 revealed the ratio between viral load and viral titer can be five orders of magnitude higher during early infection than late infection.

## MAIN TEXT

### Introduction

Throughout the COVID-19 pandemic, the relationship between the detection of viral RNA and replication-competent virus has been used as guiding evidence for infection-control strategies. For example, studies suggesting that low viral load specimens are unlikely to have observable replication-competent virus [1] were used to argue that low-analytical-sensitivity antigen tests (which only detect high viral loads [2]) would more specifically identify infectious individuals [3, 4]. Additionally, the lack of replication-competent virus in specimens collected more than a week after symptom onset [5-10] was used as evidence to release individuals from isolation despite persistently detectable viral RNA [11].

Assessment of replication-competent virus in clinical specimens is technically challenging [12] and therefore not routinely performed to determine whether an individual is infectious. Rather, the studies which have generated viral-culture data are often applied broadly to guide infection-control strategies [13]. However, the design of such studies influences the data, conclusions, and resulting policies.

Many studies that assess presence of replication-competent virus in specimens from individuals with SARS-CoV-2 infection are primarily cross-sectional, include data from only one specimen type, and are biased toward specimens collected late in the course of infection (e.g. after symptom onset) [4, 14-18]. However, during the earliest phase of infection, detection of infected individuals can help reduce subsequent transmission [19, 20] and improve clinical outcomes [21]. Few studies report viral loads starting from the incidence of acute SARS-CoV-2 infection [13, 22-29], and of these, few report both viral-load and viral-culture data [25, 27]. If studies of replication-competent virus during SARS-CoV-2 infection are insufficiently representative of early infection, resulting infection control policies may not be optimally effective.

As part of the Caltech COVID-19 Study [23, 24, 30], we attempted to fill this gap by capturing both viral load and viral titer measurements longitudinally from the incidence of acute SARS-CoV-2 infection in a subset of participants at risk of becoming infected. Within this subset, one individual was found to have incident infection with the B.1.243 lineage of SARS-CoV-2 while enrolled and collecting twice-daily specimens, from which we measured both anterior-nares (nasal) swab viral load and viral titer. This participant also collected saliva specimens for viral-load measurements. SARS-CoV-2 *N* gene viral loads and human *RNaseP* marker Cq values in saliva and nasal swab specimens from this individual (Participant AC) have previously been reported [30]. Here, we provide additional quantifications of SARS-CoV-2 *E* and *RdRp* gene viral loads and viral-titer measurements from this participant’s nasal-swab specimens to investigate the relationship of RNA viral load and infectious virus longitudinally from the incidence of naturally acquired infection.

## Results

We report the case (**Figure 1A**) of a 30–39-year-old male (Participant AC), who does not smoke/vape and is otherwise healthy (no chronic medical conditions and self-reported health as “very good”). The participant did not report evidence of prior SARS-CoV-2 infection nor receipt of any SARS-CoV-2 vaccine doses. The participant reported taking Vitamin C and fish oil supplements, and no other medications. In late-January 2021, six days prior to enrollment in this study, the participant reported exposure to SARS-CoV-2. Three days prior to enrollment, the participant began experiencing a sore throat, but two days prior to enrollment tested negative on an outpatient, non-rapid nasopharyngeal test. At this time, a household contact of Participant AC (Participant AB, **Figure S1**) tested positive, prompting eligibility of both Participant AC for enrollment in this study.

**Figure 1.**
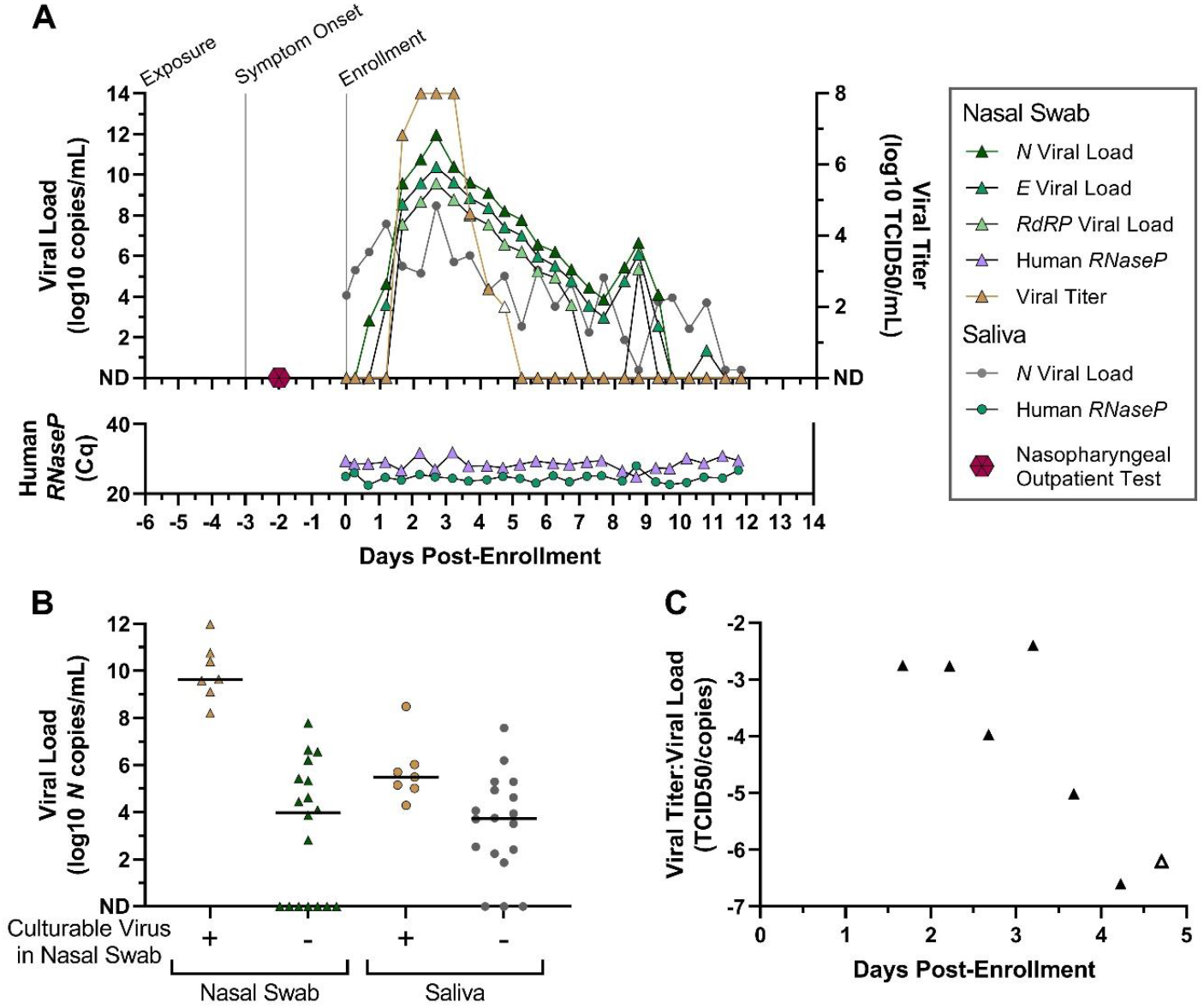
The viral load and viral titer trajectories from a single study participant from the incidence of infection. **(A)** A timeline of Participant AC’s infection is shown with notable case events (exposure, symptom onset, study enrollment), as well as SARS-CoV-2 viral loads in saliva (circles) and anterior-nares nasal swabs (triangles) on the left y-axis, and SARS-CoV-2 viral titer (log_10_ TCID_50_/mL) on the right y-axis. Human *RNaseP* Cq values are shown as a measure of sampling consistency and specimen RNA integrity. **(B)** Cross-sectional relationship of SARS-CoV-2 viral load (log_10_ *N* copies/mL, y axis) in nasal swab specimens (triangles) or saliva specimens (circles) based on whether viral culture positivity (yellow) of the nasal swab from the same timepoint. Black horizontal bars indicate median viral load. **(C)** For specimens with detectable viral titer and viral load, the ratio of viral titer (TCID_50_/mL) over *N* gene viral load (copies/mL) in nasal swab specimens collected by the participant is plotted through days of enrollment. The open symbol indicates a specimen with detectable but not quantifiable viral titer, for which 100 TCID_50_/mL was imputed. ND, not detected.

Upon enrollment, Participant AC had detectable and rising salivary viral loads, but was negative in anterior-nares nasal-swab specimens collected over the next day. During this time, the participant remained symptomatic with only a sore throat. In the subsequent day the participant developed shortness of breath and low (<10^5^copies/mL) nasal viral loads without replication-competent virus detected by culture. After this point, the participant’s nasal swab specimens achieved high (>10^7^ copies/mL) viral loads and high (>10^6^ TCID50/mL) viral titers for approximately 3 days before gradually declining. Throughout this time, headaches, cough, congestion, change in taste/smell, muscle aches, and one event of severe nausea were reported, all of which resolved before completion of enrollment.

Cross-sectional SARS-CoV-2 viral loads from different gene targets in nasal swab specimens correlated closely with each other **(Figure 1A, Figure S2A)** and the relationship between viral loads from different gene targets remained proportional throughout the course of infection **(Figure S2B)**. Cross-sectional analysis of viral load and viral titer revealed that only high viral load nasal swab specimens (>10^8^ *N* cp/mL) would contain replication competent virus (**Figure 1B**). Additionally, saliva viral load is less distinguishable between samples with and without replication competent virus in nasal swab specimens (**Figure 1B**). However, longitudinal analysis revealed that the ratio of nasal swab viral load and viral titer changed by over five orders of magnitude throughout the course of acute infection **(Figure 1C)**. This relationship indicates that RNA viral load alone, without considering infection stage, may not represent whether a specimen or a person is likely to be infectious or not.

## Discussion

High-frequency nasal swab and saliva sampling from the incidence of infection, and paired measurements of viral load and viral titer in nasal swab specimens revealed four key findings uniquely enabled by this study design.

First, saliva exhibited higher *N* gene viral loads than in nasal swabs for approximately the first two days of incident infection, after which nasal swab viral loads rose and remained subsequently higher than saliva viral loads. This supports previous observations that SARS-CoV-2 often presents first in oral specimen types before anterior nares swabs [23, 24], and that testing a single specimen type (e.g. nasal swabs) may yield false negative results during early infection.

Second, replication-competent virus was observed in nasal swabs at many timepoints when saliva viral loads were low. This suggests that the low viral load of one specimen type is not necessarily indicative of the absence of replication competent virus in another specimen type.

Third, nasal-swab viral-load measurements from different gene targets (*N, E*, and *RdRP* genes) correlated strongly with each other longitudinally, such that measurement of any one viral RNA target was indicative of other viral RNA targets [31].

Fourth, we note that the ratio between RNA viral load and culturable viral titer in nasal swabs decreased substantially (greater than 5 orders of magnitude) through the first week of infection. Cross-sectional analyses of data from participant AC and in other studies [4, 15, 18, 25, 32] have suggested a correlation between viral load and the presence of infectious virus. However, these cross-sectional analyses overlook that the relationship between viral load and infectious virus is dynamic, and that early viral loads are more indicative of viral titer than viral loads later in the infection. Therefore, earlier in the infection, individuals with lower viral loads could actually be more infectious than expected based on cross-sectional data.

Data from a SARS-CoV-2 human challenge study [25] supported these conclusions (Figure S4). In that study, 36 human participants were inoculated intranasally with 10 TCID_50_ virus, and 18 participants had subsequent sustained detectable infection. We reanalyzed longitudinal nasal swab viral load and viral culture data graciously provided by the study authors to compare to what was observed in Participant AC’s naturally acquired infection. Indeed, among specimens with replication competent virus, the average ratio between viral titer and viral load at each timepoint after inoculation decreased by nearly four orders of magnitude in the five days following inoculation.

Taken together, these results caution against conclusions about infectiousness that assume a constant ratio of RNA viral load and culturable viral titer, commonly inferred based on cross-sectional data or from single specimen types [4, 33-35]. Assuming a constant ratio of RNA viral load and culturable viral titer may not reflect early infection or all anatomical sites from which transmissible virus can be shed, and therefore may be suboptimal evidence for public health policies that seek to reduce transmission.

We acknowledge three main limitations. First, data are from a single unvaccinated person with acute SARS-CoV-2 B.1.243 infection, prior to the availability of COVID-19 vaccines and the emergence of currently circulating variants. Infection characteristics may exhibit substantial person-to-person variation, and vaccination status and/or viral variant may affect the relationship between viral load and viral titer [36]. Second, Participant AC collected saliva specimens in a preservation buffer that precluded the ability to perform viral culture, thereby prohibiting inferences on the relationship between saliva viral load and viral titer, or saliva viral titer and nasal viral titer. Third, the lack of detection of replication-competent virus by viral culture may not reflect a true absence of replication-competent virus in the specimen or shedding of infectious virus by the individual as specimen collection, handling, and storage affect virion viability [37, 38]. Moreover, both the methods of attempted viral culture and viral characteristics can affect the analytical sensitivity to detect replication-competent virus [39]. Therefore, it is possible that replication-competent virus was present in the first two nasal-swab specimens with detectable viral RNA collected by this participant, but at a concentration below the limit of detection by viral culture.

The data presented here is rare and challenging to obtain. We hope that similar datasets of viral load and viral titer in paired specimen types collected longitudinally starting from early infection can be made accessible for metanalysis and guide optimized public health strategies that reduce the burden of SARS-CoV-2 or other pathogens.

## Methods

### Participant eligibility and enrollment

Participants were recruited for a COVID-19 household transmission study under Caltech IRB protocol #20-1026 as previously described [23, 30]. All adult participants provided written informed consent.

### Study design and specimen collection

Enrolled participants began self-collecting saliva and nasal swab specimens immediately upon receipt of specimen collection materials at enrollment, and then each subsequent morning (immediately after waking), and evening (prior to bed). Participants self-collected anterior-nares nasal swabs in Nest viral transport medium (VTM) (catalog no. NST-NST-202117; Stellar Scientific, Baltimore, MD) and saliva specimens in the Spectrum SDNA-1000 saliva collection kit (Spectrum Solutions LLC, Draper, UT). Study participants were instructed not to eat, smoke, chew gum, or brush their teeth for at least 30 min prior to collection and asked to gently blow their noses before nasal swabbing (four complete rotations with gentle pressure in each nostril) with sterile flocked swabs. Specimens were transported daily by medical courier to the Caltech laboratory for analysis.

### Nucleic acid extraction, quantification of viral load by RT-qPCR, and viral variant determination

Nucleic-acid extraction was performed as previously described [23]. Conversion from RT-qPCR Cq to viral load (in copies/mL) was determined via calibration curves, reported for *N* gene previously [23], and built for *E* and *RdRP* gene using standard positive controls (IDT 10006896, IDT 10006897):

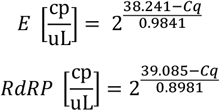

Nucleic acids extracted from the 7th saliva and nasal swab specimens collected by the participant underwent viral sequencing and variant determination as previously described [23].

### Measurement of viral titer

Tissue culture infection dose to infect 50% of test cultures (TCID_50_) assay was performed to measure the viral titer in VTM samples. Briefly, 500 μl VTM sample was filter-cleaned with a spin column (CLS-8160, Corning). VeroE6 cells ectopically expressing human ACE2 and TMPRSS2 (VeroE6-AT cells; a gift from Dr. Barney Graham, National Institutes of Health, Bethesda MD) were seeded confluent in a 96-well plate, after replacing the seeding medium with 90 µL of assay medium (Dulbecco’s Modified Eagle Media (DMEM) + 2% heat inactivated Fetal Bovine Serum (FBS) + 10 mM HEPES + 1% Penicillin/Streptomycin), 10 µL of filtered VTM sample was added to the first row of the plate as the starting inoculation. Then, 10-fold serial dilutions were performed in the 2nd through 7th rows, leaving the 8th row as the negative control. Each sample was tested with 5 replicates. Cells were fixed with 10% formaldehyde and stained with 1% crystal violet three days post infection. Digital photographs were taken, and cell death indicated by clear areas in a well, were scored to calculate TCID_50_.

## Data Availability

The data underlying the results presented in the study are available at CaltechDATA at https://data.caltech.edu/records/cgf4q-byr92.

## Acknowledgements

We wish to thank the participant who contributed these specimens for analysis, the contact tracers at the Pasadena Public Health Department, as well as Dr. Matthew Bidwell Goetz and Dr. David Beehouwer for their thoughts on this data. We thank Maira Phelps, Lienna Chan, Lucy Li, Dan Lu, and Amy Kistler at the Chan Zuckerberg Biohub for performing SARS-CoV-2 sequencing.

