## Supplemental Information for "The ratio between SARS-CoV-2 RNA viral load and culturable viral titer differs depending on stage of infection"

**Contents**

Table S1

Figures S1-S3

Detailed Author Contribution Statements

**Table S1. Reagent List.** Table includes all reagents utilized in this study.

| Step | ReagentName | Description | Manufacturer | Catalogue Number |
|---|---|---|---|---|
| Specimen Collection | Spectrum SDNA1000 Saliva Collection Device | For at-home collection of spit saliva into a guandinium-thiocyanate based preservation buffer | Spectrum Solutions LLC | SDNA1000 |
| Specimen Collection | NEST Scientific 10mL Sterile Screw-Cap Transport Tube with 3mL VTM | For at-home collection of nasal swab specimens into media that maintains live virions | Stellar Scientific | NST-NST-202117 |
| Nucleic Acid Extraction | MagMAX™ Viral/Pathogen Nucleic Acid Isolation Kit | For extraction of nucleic acids from clinical upper respiratory specimens | ThermoFisher Scientitif | A42352 |
| Viral Load Quantification | TaqPath™ COVID-19 Combo Kit | For RT-qPCR measurement of human RNaseP and SARS-CoV-2 N gene | ThermoFisher Scientific | A47814 |
| Viral Load Quantification | Heat-inactivated SARS-CoV-2 2019-nCoV/USA-WA1/2020 | Extraction control and standard for RT-qPCR quantification | BEI | NR-52286 |
| Viral Load Quantification | 2019-nCoV_E Positive Control | Standard for RT-qPCR quantification | IDT | 10006896 |
| Viral Load Quantification | 2019-nCoV_RdRp (ORF1ab) Positive Control | Standard for RT-qPCR quantification | IDT | 10006897 |
| Viral Load Quantification | E_Sarbeco_F1 Forward Primer, 50 nmol | Forward primer for RT-qPCR measurement of SARS-CoV-2 E gene | IDT | 10006888 |
| Viral Load Quantification | E_Sarbeco_R2 Reverse Primer, 50 nmol | Reverse primer for RT-qPCR measurement of SARS-CoV-2 E gene | IDT | 10006890 |
| Viral Load Quantification | E_Sarbeco_P1 (FAM) Probe, 25 nmol | Probe for RT-qPCR measurement of SARS-CoV-2 E gene | IDT | 10006892 |
| Viral Load Quantification | RdRP_SARSr_F2 Forward Primer, 50 nmol | Forward primer for RT-qPCR measurement of SARS-CoV-2 RdRp gene | IDT | 10006860 |
| Viral Load Quantification | RdRP_SARSr_R1 Reverse Primer, 50 nmol | Reverse primer for RT-qPCR measurement of SARS-CoV-2 RdRp gene | IDT | 10006881 |

| | | | | |
|---|---|---|---|---|
| Viral Load Quantification | RdRP_SARSr_P2 (SUN) Probe, 25 nmol | Probe for RT-qPCR measurement of SARS-CoV-2 RdRp gene | IDT | 10007063 |
| Viral Culture | Culture Media | DMEM<br>2%FBS (heat inactivated)<br>1% Penicillin-Streptomycin<br>1% HEPES (1M) | Fisher Scientific<br>Fisher Scientific | MT10013CV<br>SH30071.03<br>MT30002CI<br>MT25060CI |
| Viral Culture | Cell Line | VeroE6-AT | A gift from Dr. Barney Graham (NIH) | |
| Viral Culture | Stain for Readout | 1% crystal violet<br>20% Ethanol | Sigma_Aldrich<br>Fisher | C-6158<br>4355222 |

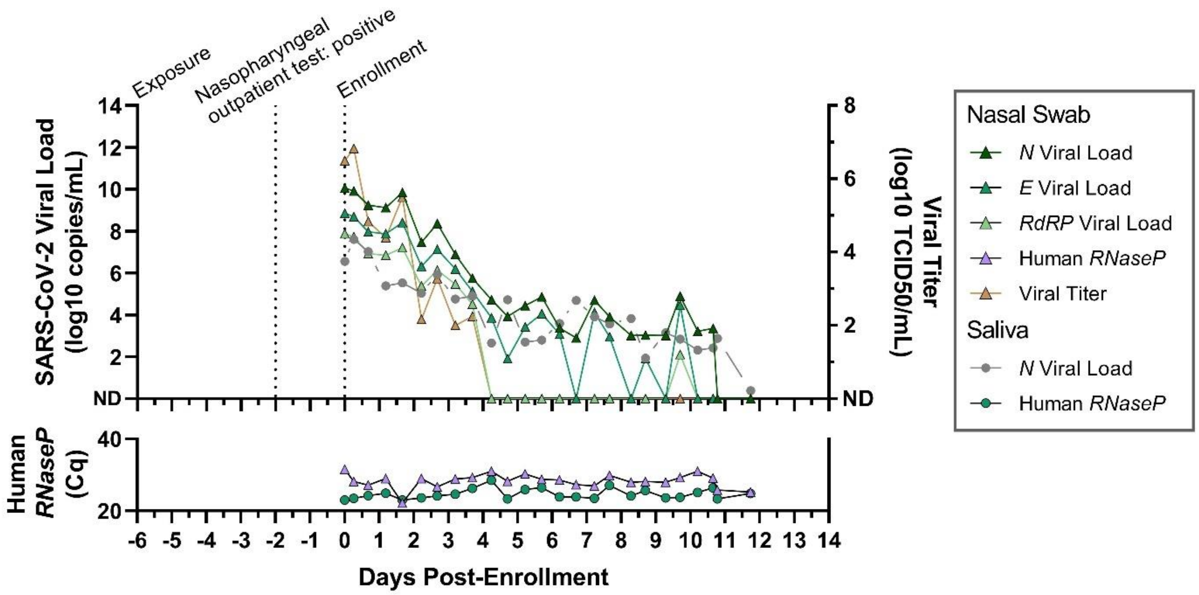


**Figure S1. The viral load and viral titer trajectories from a single study participant from the incidence of infection. (A)** A timeline of this participant's infection is shown with notable case events (e.g. exposure, positive nasopharyngeal outpatient test, study enrollment), as well as SARS-CoV-2 viral loads (log10 copies/mL) in saliva (circles) and anterior nares nasal swab (triangles) on the left y-axis, and SARS-CoV-2 viral titer (log10 TCID50/mL) on the right y-axis. Human *RNaseP* Cq values are shown as a measure of sampling consistency and specimen RNA integrity. ND, not detected.

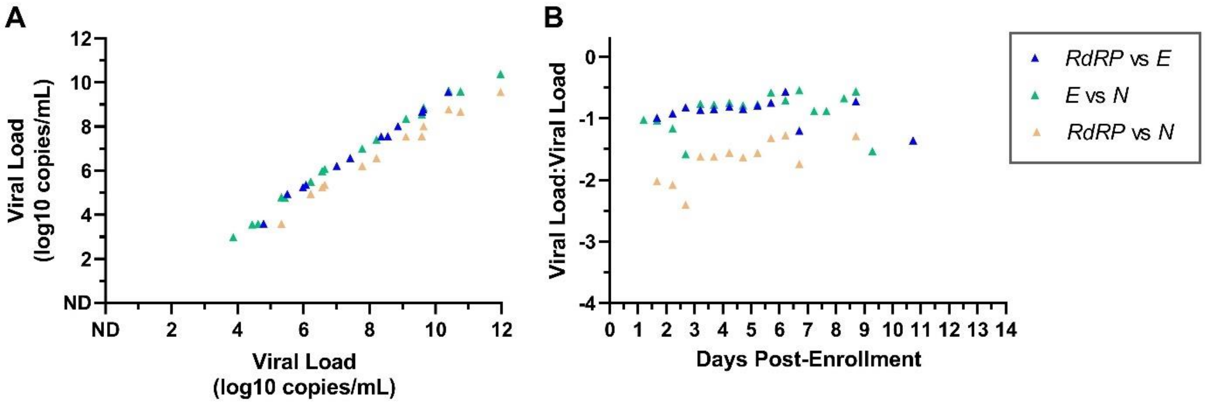


**Figure S2. Swab viral loads measured from *N*, *E*, and *RdRP* genes remain constant with respect to each other through the course of infection. (A)** The viral load from one gene is plotted on the y axis with respect to another gene comparing *RdRP* and *E* genes (blue triangle), *E* and *N* genes (green triangles), and *RdRP* and *N* genes (tan triangles). **(B)** The ratios of viral loads are plotted over days post-enrollment for RdRP and *E* genes (blue triangle), *E* and *N* genes (green triangles), and *RdRP* and *N* genes (tan triangles). Viral loads that were not detected were omitted from analysis. ND, not detected.

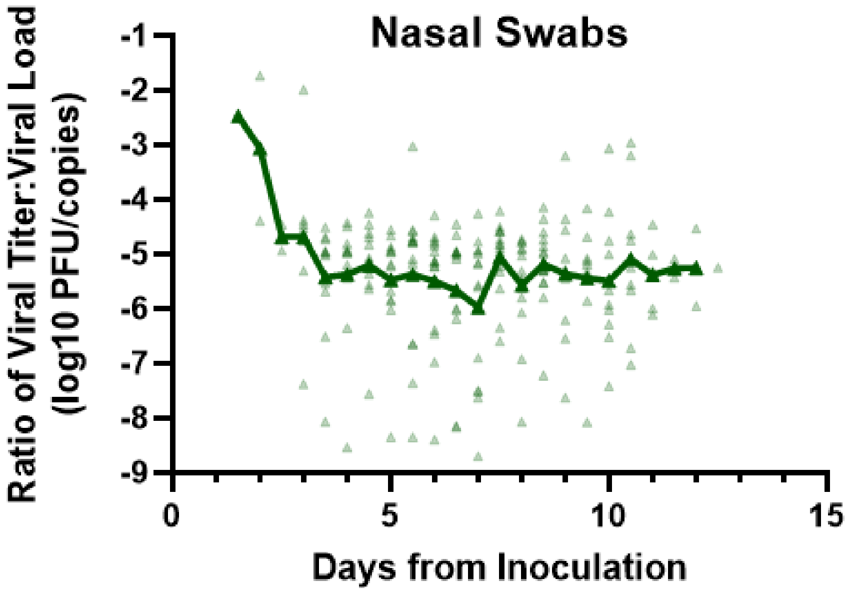


**Figure S3. Longitudinal Ratio of Viral Titer to Viral Load from Participants in SARS-CoV-2 Human Challenge Study.** As part of a SARS-CoV-2 human challenge study performed in (cite Killingly) participants were inoculated intranasally with 10 $TCID_{50}$ virus. 18 participants had subsequent sustained detectable infection in nasal swab and throat swab specimens collected daily after inoculation. Viral load and viral culture data from these specimens was graciously provided by the authors of this study. We plotted the log10 transformed ratio of viral titer to viral load in nasal swabs, for all specimens with replication competent virus, by the time from inoculation (green triangles). Green line represents the average log10 transformed ratio of viral titer to viral load among culture-positive nasal swab specimen, for each day following inoculation.

**Author Contribution Statements:**

Michael K. Porter (MKP)- Conceptualization of nested study with AVW and RFI. Literature review with AVW. Co-wrote biosafety protocols for receipt and processing of live specimens with AVW. Reagent and supply acquisition. Determined and validated live specimen collection methods with AVW. Received, logged and aliquoted live specimens with AVW. Established academic collaboration with colleagues at University of Washington. Performed nucleic acid extractions and *RNaseP, N, E, and RdRP* gene RT-qPCR. Coordinated and prepared shipments of live specimens to collaborators. Interpreted paired viral load and culture data with AVW and LH. Outlined and co-wrote manuscript with AVW. Prepared Figures 1, S1, S2, and S3. Validated underlying data with AVW. Managed author citations and references.

Alexander Viloria Winnett (AVW)- Conceptualization of nested study with MKP and RFI. Literature review with MKP. Co-wrote biosafety protocols for receipt and processing of live specimens with MKP. Reagent and supply acquisition. Determined and validated live specimen collection methods with MKP. Received, logged and aliquoted live specimens with MKP. Performed nucleic acid extractions and *RNaseP* and *N* gene RT-qPCR. Coordinated and prepared shipments of nucleic acids for viral sequencing. Coordinated and prepared shipments of live specimens to collaborators for viral culture. Interpreted paired viral load and culture data with MKP and LH. Outlined and co-wrote manuscript with MKP. Provided feedback on the design of Figures 1, S1, and S2. Validated underlying data with MKP.

Linhui Hao (LH)- Received live specimens for viral culture. Performed viral culture at BSL3 and analyzed viral culture data. Interpreted paired viral load and culture data with AVW and MKP. Wrote viral culture methods section. Edited the manuscript.

Natasha Shelby (NS)- Study Administrator; collaborated on study design and recruitment strategies; created instructions for live viral-load sampling with JAR; enrolled and maintained study participants with JAR and NS. Edited the manuscript.

Jessica A. Reyes (JAR)- Lead Study Coordinator; created instructions for live viral-load sampling with NS; enrolled and maintained study participants with NS and NWS.

Noah W. Schlenker (NWS)- Study Coordinator; enrolled and maintained study participants with NS and JAR.

Anna E. Romano (AER)- Analyzed and summarized viral sequencing data.

Colton Tognazzini (CT)- Coordinated the recruitment efforts at PPHD with case investigators and contact tracers and provided guidance and expertise on SARS-CoV-2 epidemiology and local trends.

Matthew Feaster (MF)- Co-investigator; collaborated on study design and recruitment strategies; provided guidance and expertise on SARS-CoV-2 epidemiology and local trends.

Ying-Ying Goh (YYG)- Co-investigator; collaborated on study design and recruitment strategies; provided guidance and expertise on SARS-CoV-2 epidemiology and local trends.

Michael Gale (MG)- Co-investigator; coordinated and oversaw viral culture method development and execution. Edited the manuscript.

Rustem F. Ismagilov (RFI)- Co-investigator; collaborated on study design and recruitment strategies; provided leadership, technical guidance, oversight, and was responsible for obtaining funding for the study.
